# Birth Weight and Associated Factors among Host and Refugee Neonates at Health Facilities in Gambella Region: A facility based Comparative Cross-sectional Study

**DOI:** 10.1101/2022.06.27.22276951

**Authors:** Bang Chuol Nhial, Shambel Wedajo, Sisay Eshete Tadesse

## Abstract

**Background:** Birth weight remains as one of the facing factors and one of the leading causes of child suffering worldwide. This is a major problem especially in low and middle income countries and most importantly in vulnerable populations like refugee. However, there is a limited evident as yet in the study area.

**Objective:** The aim of this study was to assess the birth weight and associated factors among host and refugee neonates at health facilities in Gambella Region.

**Methods:** A facility based comparative cross-sectional study was applied in host and refugee settings. A total of five hundred ninety six neonates were included in this study from February 1^st^, 2020 to March 31^st^, 2020 through multi-stages sampling technique. The data were collected using structured interview and documents review methods with structured questionnaires as instruments. Pretesting of the tool and training of data collectors and field supervision were made. Using statistical package for social science version 25, variables with p≤0.05 from multiple linear regression analysis were declared as factors significantly associated with the birth weight. Furthermore, independent samples t-test was computed to compare the mean birth weights.

**Results:** The study resulted in 100% response rate. The mean birth weights were found to be significantly difference between host and refugee neonates with means of 3282.55±415.97 grams and 3109.40±635.10 grams respectively (m_1_-m_2_ = 173.15, 95% CI: (86.75, 259.56)). Several factors such as number of fetuses (β = -148.35, 95% CI: (-234.86, -61.83)), gestational age at delivery (β = 90.83, 95% CI: (66.72, 114.93)), household food security status (β = 166.33, 95% CI: (100.68, 231.97)), individual dietary diversity score (β = 88.75, 95% CI: (68.69, 108.81)) and pregnancy induced hypertension (β = -148.35, 95% CI: (-234.86, -61.83)) were found to be the most influential factors significantly associated with the birth weight.

**Conclusions:** In the study area, host neonates have larger mean birth weight than the refugee neonates. Number of fetuses, gestational age at delivery, household food security, individual dietary diversity score and pregnancy induced hypertension were factors influencing the birth weight.

## Background

According to the world health organization (WHO), birth weight (BW) is defined as the first weight of an infant measured within first hour after birth before significant occurrence of postnatal weight lost. When it is found to be less than 2500 grams (up to and including 2499 grams) irrespective of the gestational age of the neonate it can be termed as low birth weight (1). Birth weight (BW) determines the child’s probability to survive as well as susceptibility to various risks of childhood morbidities (2,3). According to United Nation Children’s Fund (UNICEF) and World Health Organization (WHO), birth weight can be classified in to high birth weight, normal birth weight, low birth weight, very low birth weight and extremely low birth weight (3).

An approximate of 3.6 million deaths of infants occurred during the neonatal period is accounted to birth weight problem that is nearly 60% to 80% (4).

More than 20 million infants worldwide representing 15.5% of all births are born with low birth weight, 95.6% of them are found in developing countries (3,4). The level of low birth weight in developing countries (16.5%) is more than twofold the level in developed regions (3,5). Half of all low birth weight babies are born in South-central Asia, where more than a quarter (27%) of all infants weight less than 2,500 grams at birth (5). A low birth weight level in Saharan Africa is around 15% (4). People of concern to United Nation High Commissioner for Refugee (UNHCR) were estimated to be 42.5 million. Of these, refugees accounted 15.2 million, internally displaced persons (IDPs) 26.4 million and other individuals whose asylum applications had not succeeded constituted for 895,000 (6).

Of all neonatal deaths, more than a half of them occur in countries with neonatal mortality rate of 30 and above deaths per 1000 live births. Most of these countries have experienced recent conflict and residing for refuge in the other countries in seeking for humanitarian aid (7). An estimate of LBW includes 28% in South Asia, 13% in Saharan Africa and 9% in Latin America. The incidence of LBW also increases at alarming rate in developing countries like Ethiopia that is why LBW has drawn attention as public health issue (8,9). The WHO countries cooperation strategy 2008–2011showed that the prevalence of low birth weight in Ethiopia is 14%, which is one of the highest in the world (10).

The magnitude of low birth weight is going on increasing as time goes (8).

The problem of birth weight is not only a leading cause of childhood mortality and morbidity but also the causative agent for increment of risk for non-communicable diseases (NCD) later in life (2,11,12). Evidence indicated that low birth weight infants are 2 times more likely to die during the first 28 days of their live compared with the normal ones (4). This is a major problem especially in low and middle income countries and in the vulnerable populations like refugees. Birth weight problem varies across countries and within country.

The world health Assembly (WHA) Resolution 65.6 authorized a complete implementation plan on maternal, infant and young child nutrition aiming at reducing 30% of LBW with the most purpose of enhancing attention to, investment in and action for a group of cost effective intervention and policies which will help the member states and their partners in lowering the rate of LBW by the year 2025 (13). The aim of LBW decrement by at least one third between the year 2000 and 2010 was one among the major goals in ‘A World Fit for Children’, the Declaration and Plan of Action adopted by the United Nations General Assembly session on Children in the year 2002 (13).

Though there were many studies conducted in Ethiopia, there is a limited evident as yet in the study area. For example a few studies (if any) were done aimed at assessing the birth weight and associated factors among host and refugee neonates simultaneously following appropriate model for the outcome variable and a few (if any) were conducted in Gambella Region. So the aim of this study was to assess the birth weight and associated factors among host and refugee neonates at health facilities in Gambella Region following comparative cross-sectional study design with multiple linear regression model.

## Methods

### Study area

This study was conducted in Gambella Regional State which is one of the ten regional states of Ethiopia with its capital city Gambella. The region is about 766 km away from Ethiopia’s capital city, Addis Ababa. The region is located in South West of the country and bordered with the Oromiya Regional State in the East; with Benishangul-Gumuz Regional State in the North; with the Southern Nations Nationalities and People’s Regional State in the South and East; with the Republic of South Sudan in the West and North. On the basis of Ethiopia census conducted by Central Statistical Agency (CSA) in 2007, the Gambela Region had an approximate population of 307,096, comprising of 159,787 males and 147,309 females with urban occupants of 77,925 or 25.37% of the population (14). The region has fourteen woredas and seven camps hosting mostly South Sudanese with approximate total refugee population of 304447 (14).

### Study design and period

A facility based comparative cross-sectional study design was conducted from February 1^st^, 2020 up to March 31^st^, 2020 in the selected health facilities in the host and refugee settings in Gambella Region.

### Source population

All neonates born in health facilities in the host and refugee settings in Gambella Region

### Study population

All neonates born in the selected health facilities in the host and refugee settings in Gambella Region during the time of data collection

### Study variables

The dependent variable was the birth weight whereas independent variables include socio-demographic factors (neonatal sex, maternal age, marital status, house ownership, maternal educational status, partner educational status, maternal occupational status, partner occupational status, family size and household monthly income), maternal factors (number of fetuses, parity, age at first birth, gestational age at delivery, working time and number of ANC visits), nutrition related factors (dietary counseling during pregnancy, number of iron tablet used during pregnancy, extra meal intake during pregnancy, household food security status, alcohol consumption during pregnancy, khat chewing habit during pregnancy, smoking habit during pregnancy and individual dietary diversity score) and health related factors of mother (pregnancy induced hypertension, malaria status during pregnancy and anemia status during pregnancy).

### Sample size determination

The sample size was calculated using double populations means comparison formula with the subsequent assumptions; 95% level of confidence and 80% power of the study. With birth weight of mean (m_1_) and standard deviation (S_1_) of 3245.3 grams and 51.8 grams respectively among the host peoples’ neonates (15,16). With birth weight of mean (m_1_) and standard deviation (S_1_) of 3100 grams and 600 grams respectively among the refugees’ neonates (16,17).

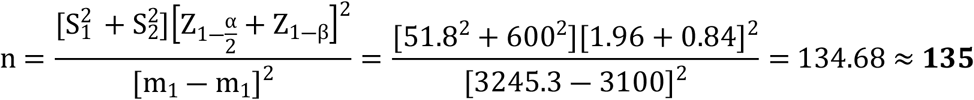

Adding 10% for possible non-response rates and considering the design effect of 2, the sample size was **298** neonates in each setting giving a final sample size of **596** neonates.

### Sampling methods

A multi-stages sampling technique was employed in selection of study settings, health facilities and neonates in Gambella Region.

### Data collection

Data were collected through face to face maternal interview using structured questionnaire and medical record review of the mothers and newborns by seven nurses of BSc holders were recruited as data collectors supervised by three BSc holders who were recruited as supervisors.

### Data quality control

Questionnaire was translated into local language (Nuer) and reviewed for the actual completion and the inclusion of all questions important for the objective of the study. Training was given to the supervisors and data collectors for two days. The questionnaire was pre-tested in one health facility that was not included in the study in each setting and minor corrections were made accordingly.

### Data processing and management

Date entry was done using Epi Info version7 and transferred into SPSS version 25 for analysis. Before the actual analysis, data were first checked for completeness and consistence.

### Data analysis

After data entry was done using Epi Info version 7 and transferred into SPSS version 25 for analysis, descriptive and inferential statistics were performed. Inferential statistics such as independent samples t-test and multiple linear regression were computed. Independent samples t-test was performed to compare the mean birth weight. Multiple linear regression model was performed to identify the factors associated with the birth weight (18).

### Ethical considerations

Ethical clearance was obtained from Ethic Review Committee (ERC) of School of Public Health, College of Medicine and Health Sciences, Wollo University. Written informed consent was obtained from each study participant and confidentiality was maintained.

## Results

This study was done on previously planned study subjects of 596 (298 from host and 298 from refugee) neonates resulting in 100% response rate. Half of neonates from host (154(51.7%)) and refugee (148(49.7%)) are males. The minimum and the maximum birth weight among host neonates are 1500 and 4300 grams respectively with mean of 3282.55±415.97 grams whereas the minimum and the maximum birth weight among refugee neonates are 1200 and 4200 grams respectively with mean of 3109.40±635.10 grams (Table 1)

**Table 1:** Socio-demographic characteristics of study participants at health facilities in Gambella Region, Ethiopia, July, 2020.

| Nominal variables |  |  |  |  |  |  |  |
| --- | --- | --- | --- | --- | --- | --- | --- |
| Variable | Category | Host<br>(n = 298) |  | Refugee<br>(n = 298) |  | Total (Both)<br>(n = 596) |  |
|  |  | Freq. | % | Freq. | % | Freq. | % |
| Neonatal sex | Male | 154 | 51.7 | 148 | 49.7 | 302 | 50.7 |
|  | Female | 144 | 48.3 | 150 | 50.3 | 294 | 49.3 |
| Marital status | Married | 273 | 91.6 | 282 | 94.6 | 555 | 93.1 |
|  | Unmarried | 25 | 8.4 | 16 | 5.4 | 41 | 6.9 |
| House ownership | Yes | 248 | 83.2 | 258 | 86.6 | 506 | 84.9 |
|  | No | 50 | 16.8 | 40 | 13.4 | 90 | 15.1 |
| Maternal educational status | Educated | 227 | 76.2 | 173 | 58.1 | 400 | 67.1 |
|  | Uneducated | 71 | 23.8 | 125 | 41.9 | 196 | 32.9 |
| Partner educational status | Educated | 234 | 78.5 | 167 | 56.0 | 401 | 67.3 |
|  | Uneducated | 64 | 21.5 | 131 | 44.0 | 195 | 32.7 |
| Maternal occupational status | Employed | 93 | 31.2 | 67 | 22.5 | 160 | 26.8 |
|  | Unemployed | 205 | 68.8 | 231 | 77.5 | 436 | 73.2 |
| Partner occupational status | Employed | 147 | 49.3 | 34 | 11.4 | 181 | 30.4 |
|  | Unemployed | 151 | 50.7 | 264 | 88.6 | 415 | 69.6 |
| Scale variables |  |  |  |  |  |  |  |
| Variable | Host |  | Refugee |  | Total (Both) |  |  |
| | Min-Max | Mean ± SD | Min-Max | Mean ± SD | Min-Max | Mean ± SD | |
| Birth weight (gram) | 1500-4300 | 3282.55 ± 415.97 | 1200-4200 | 3109.40 ± 635.10 | 1200-4300 | 3195.97 ± 543.33 | |
| Maternal age (year) | 16-38 | 24.51 ± 5.09 | 16-41 | 26.08 ± 5.64 | 16-41 | 25.30 ± 5.43 | |
| HMI (ETB) | 1500-26000 | 10257.05 ± 6101.83 | 1500-12000 | 4834.90 ± 2897.82 | 1500-26000 | 7545.97 ± 5489.88 | |

Two hundred ninety six host neonates (99.3%) and two hundred ninety five refugee neonates (99.0%) are singletons. The minimum and the maximum gestational age at delivery of host mothers are 35 and 41 weeks respectively with mean of 38.18±1.24 weeks compared to the minimum and the maximum gestational age at delivery of refugee mothers of 34 and 40 weeks respectively whose mean is 37.93±1.46 weeks The minimum and the maximum working time of host and refugee are the same and equal to 1 and 9 hours respectively. The mean of working time of host and refugee mothers are 5.44±2.00 and 5.42±1.93 hours respectively (Table 2).

**Table 2:**
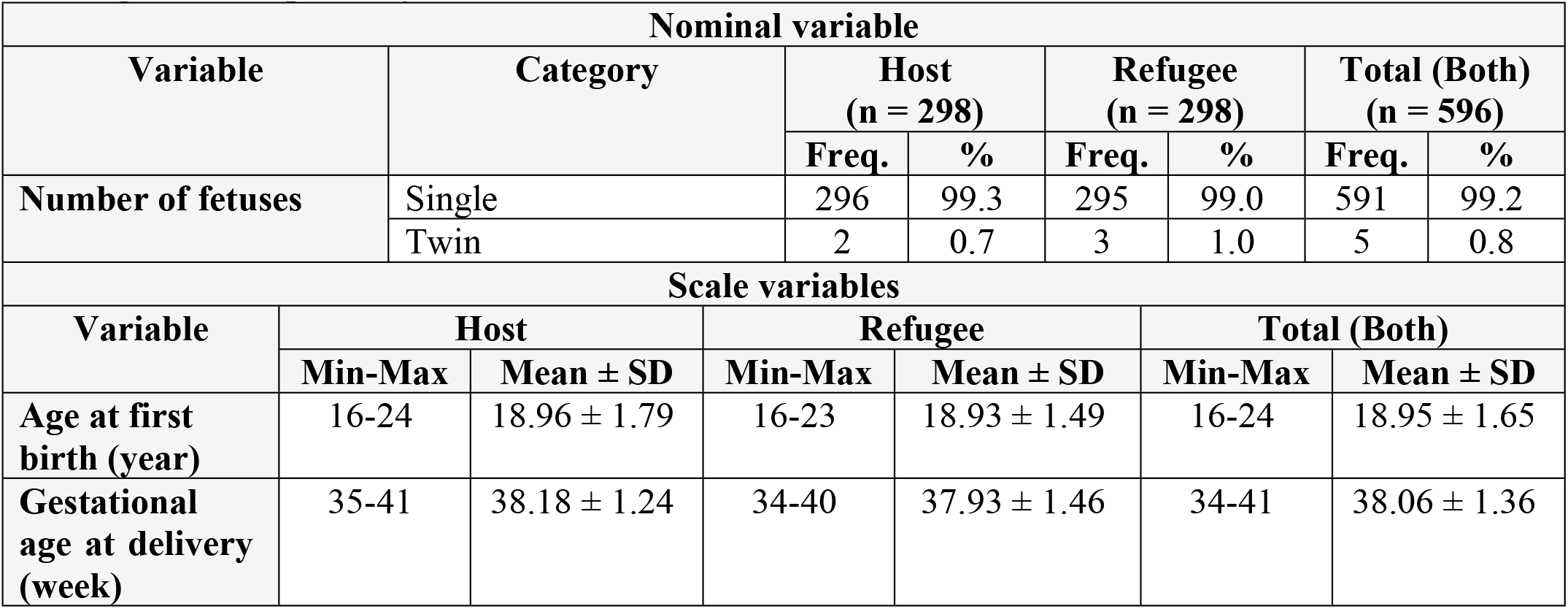
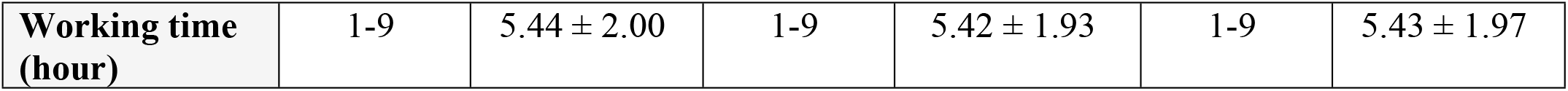
Maternal characteristics of study participants at health facilities in Gambella Region, Ethiopia, July, 2020.

| Nominal variable |  |  |  |  |  |  |  |
| --- | --- | --- | --- | --- | --- | --- | --- |
| Variable | Category | Host<br>(n = 298) |  | Refugee<br>(n = 298) |  | Total (Both)<br>(n = 596) |  |
|  |  | Freq. | % | Freq. | % | Freq. | % |
| Number of fetuses | Single | 296 | 99.3 | 295 | 99.0 | 591 | 99.2 |
|  | Twin | 2 | 0.7 | 3 | 1.0 | 5 | 0.8 |
| Scale variables |  |  |  |  |  |  |  |
| Variable | Host |  | Refugee |  | Total (Both) |  |  |
| | Min-Max | Mean ± SD | Min-Max | Mean ± SD | Min-Max | Mean ± SD | |
| Age at first birth (year) | 16-24 | 18.96 ± 1.79 | 16-23 | 18.93 ± 1.49 | 16-24 | 18.95 ± 1.65 | |
| Gestational age at delivery (week) | 35-41 | 38.18 ± 1.24 | 34-40 | 37.93 ± 1.46 | 34-41 | 38.06 ± 1.36 | |

|  |  |  |  |  |  |  |
| --- | --- | --- | --- | --- | --- | --- |
| **Working time (hour)** | 1-9 | 5.44 ± 2.00 | 1-9 | 5.42 ± 1.93 | 1-9 | 5.43 ± 1.97 |

One hundred eighty (60.4%) of mothers from host and one hundred sixty five (55.4%) of mothers from refugee had dietary counseling during pregnancy. Three fourth of mothers from host community (231(77.5%)) and less than three fourth from refugee (210(70.4%)) are food secure. Sixteen (5.4%) and thirty three (11.1%) host and refugee mothers respectively were alcohol consumers during pregnancy. Only two mothers (0.7%) in both host and refugee were khat chewers. Fifteen mothers from host community (5.0%) and twenty four mothers from refugee (8.1%) were smoking during pregnancy likewise only sixteen among host mothers (5.4%) and thirty three among refugee mothers (11.1%) were consuming alcohol during pregnancy (Table 3).

**Table 3:**
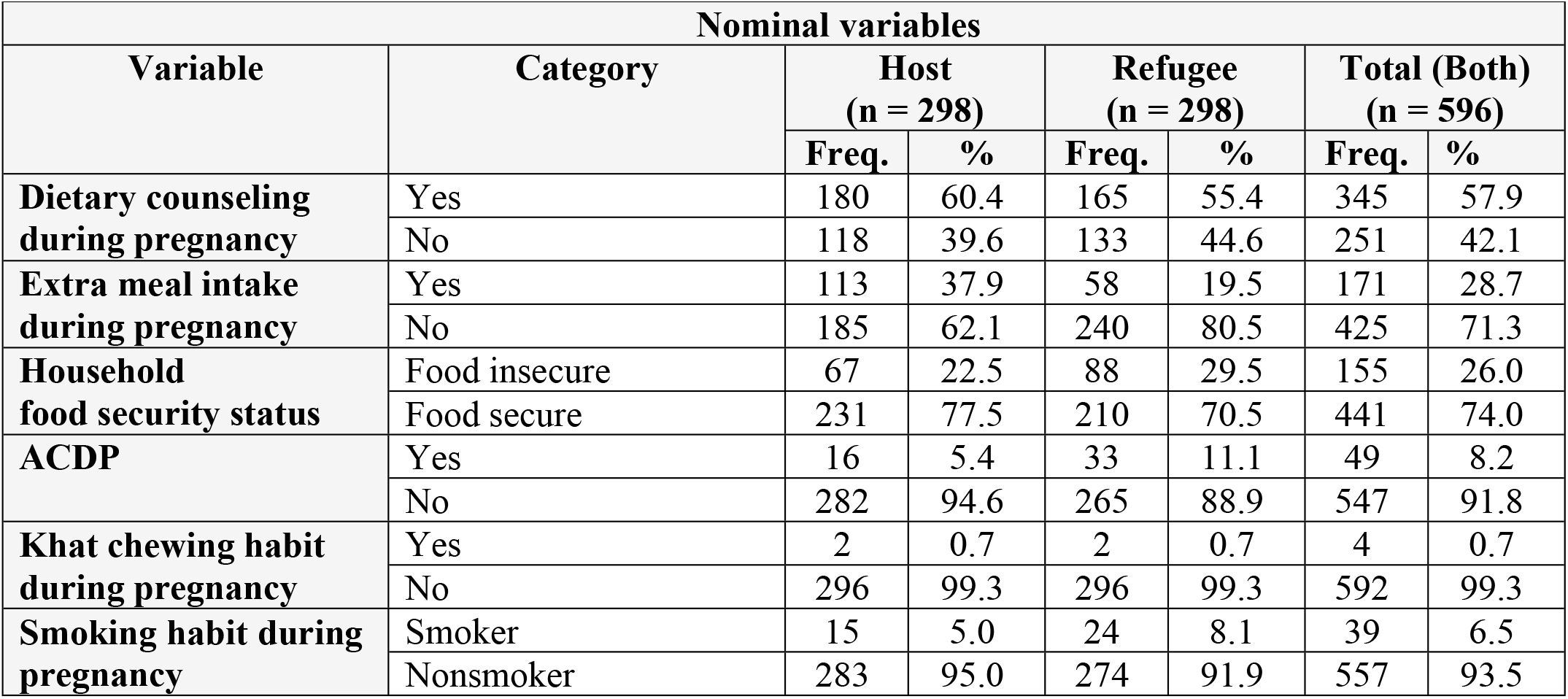
Nutrition related characteristics of study participants at health facilities in Gambella Region, Ethiopia, July, 2020.

Only twenty five mothers from host community (8.4%) and thirty eight from refugee (12.8%) had pregnancy induced hypertension during pregnancy. Less than half of mothers from host community (122(40.9%)) had malaria during pregnancy whereas exactly half of mothers from refugee (149(50.0%)) had malaria during pregnancy. Fifty mothers from host (16.8%) and ninety six mothers from refugee (32.2%) had anemia during pregnancy (Table 4).

**Table 4:** Health related characteristics of study participants at health facilities in Gambella Region, Ethiopia, July, 2020.

| Nominal variables |  |  |  |  |  |  |  |
| --- | --- | --- | --- | --- | --- | --- | --- |
| Variable | Category | Host (n = 298) |  | Refugee (n = 298) |  | Total (Both) (n = 596) |  |
|  |  | Freq. | % | Freq. | % | Freq. | % |
| **Pregnancy induced hypertension** | Yes | 25 | 8.4 | 38 | 12.8 | 63 | 10.6 |
|  | No | 273 | 91.6 | 260 | 87.2 | 533 | 89.4 |
| **Malaria status during pregnancy** | Yes | 122 | 40.9 | 149 | 50.0 | 271 | 45.5 |
|  | No | 176 | 59.1 | 149 | 50.0 | 325 | 54.5 |
| **Anemia status during** | Yes | 50 | 16.8 | 96 | 32.2 | 146 | 24.5 |
| pregnancy | No | 248 | 83.2 | 202 | 67.8 | 450 | 75.5 |

### Prevalence of low birth weight

The prevalence of low birth weight among host and refugee neonates are 1% and 17.8% respectively with overall prevalence in the region of 9.4% (95% CI: (12.3%, 21.3%)).

The host and refugee neonates have birth weights of means of 3282.55±415.97 and 3109.40±635.10 grams respectively. Hence, by considering the independent samples t-test for equality of means assuming the difference in variance, it follows that the mean birth weights of host and refugee neonates are significantly difference (µ_1_-µ_2_ = 173.15, 95% CI: (86.75, 259.56)) (Table 5).

**Table 5:**
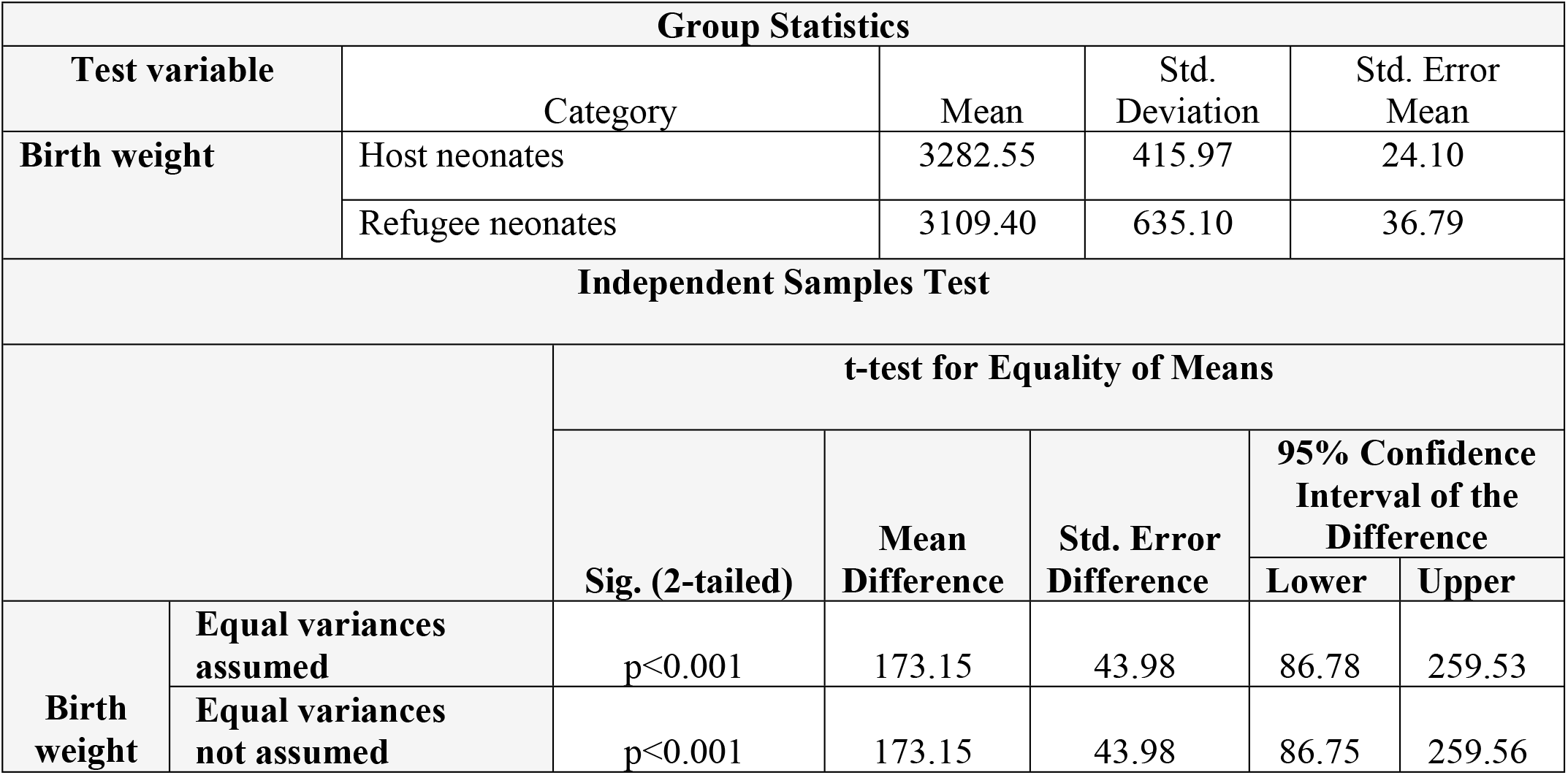
Comparison of mean birth weights between host and refugee neonates at health facilities in Gambella Region, Ethiopia, July, 2020 (n = 298)

**Table 6:**
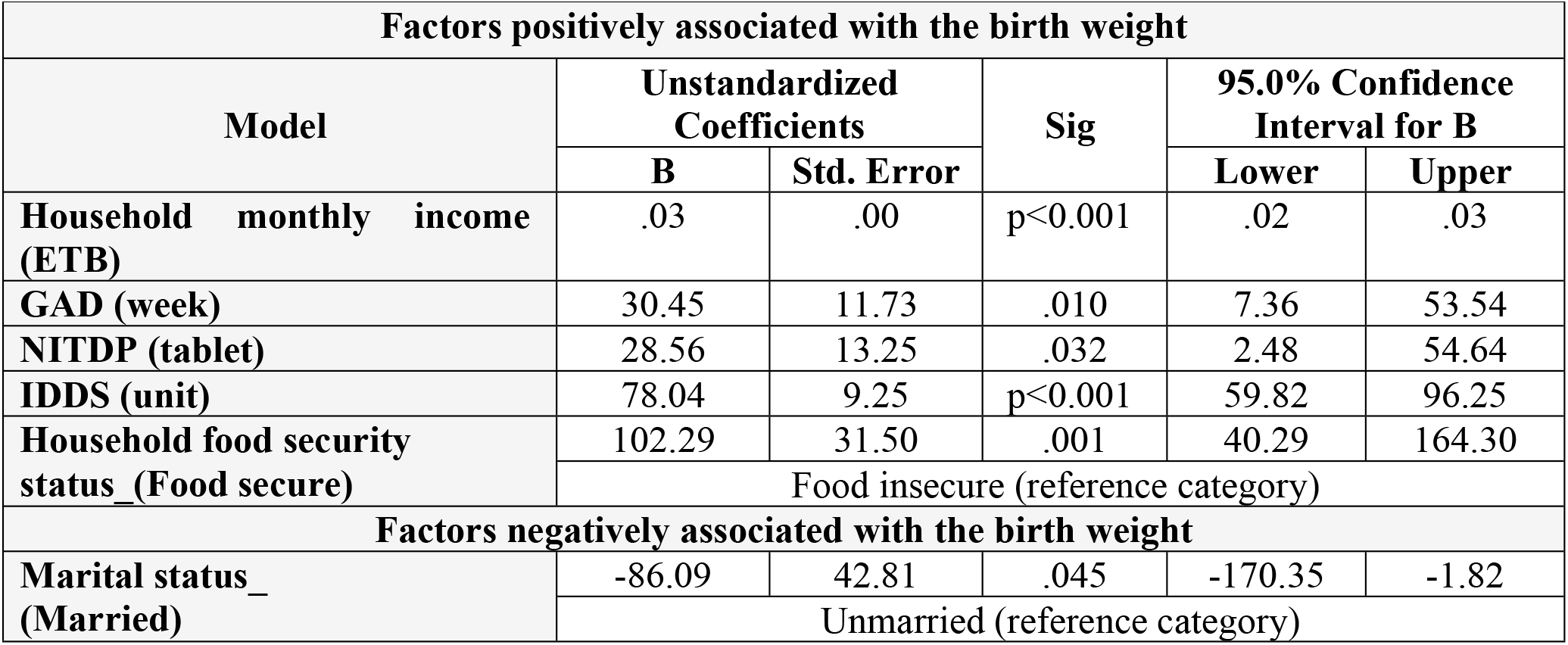
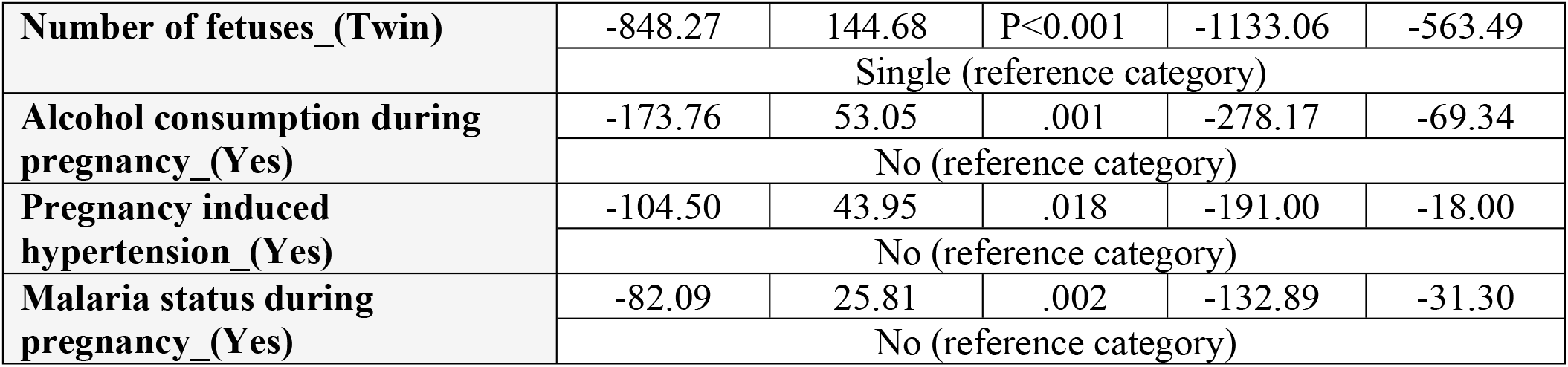
Factors associated with birth weight at host community health facilities in Gambella Region, Ethiopia, July, 2020 (n = 298)

**Table 7:**
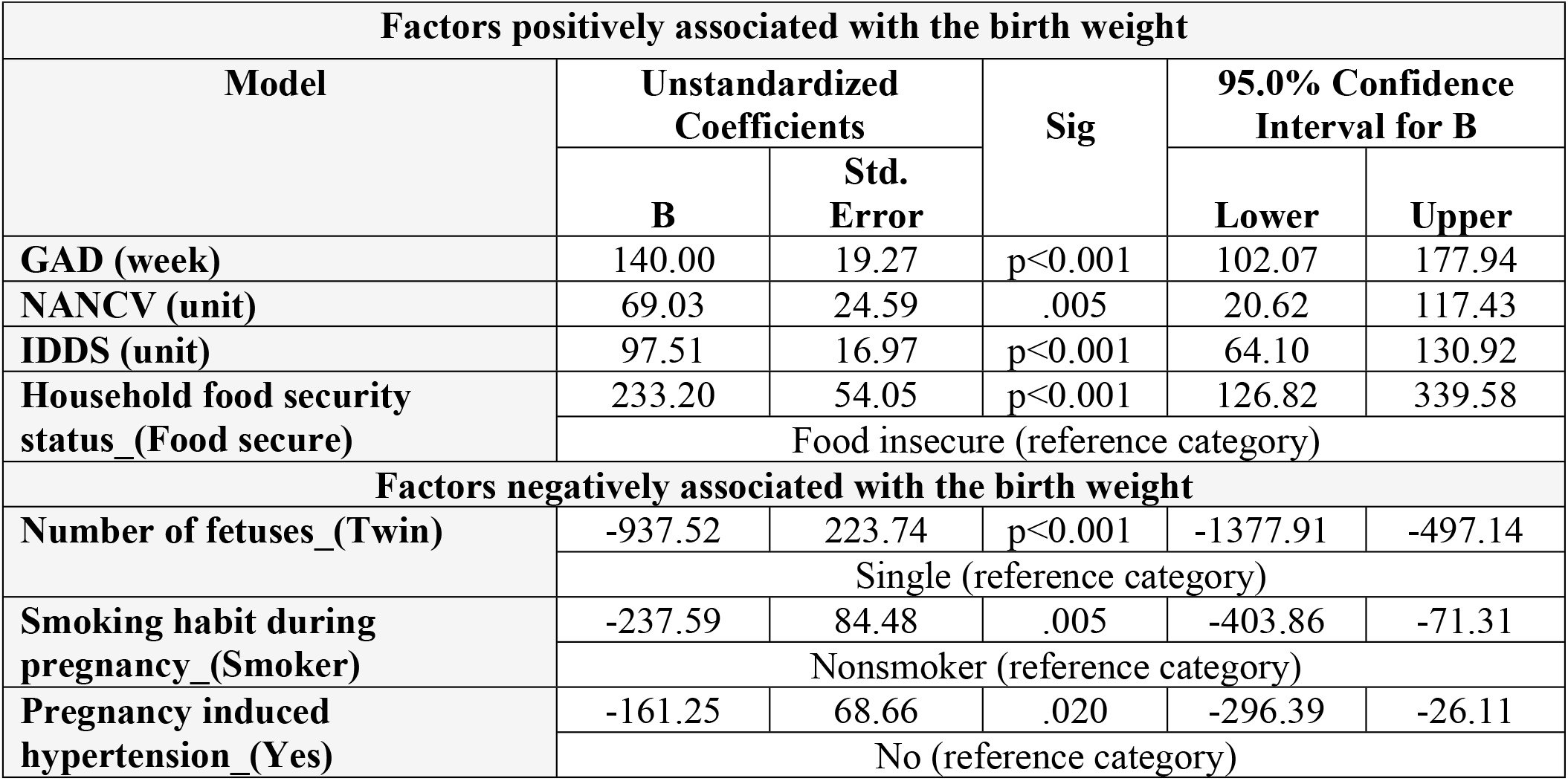
Factors associated with birth weight at refugee health facilities in Gambella Region, Ethiopia, July, 2020 (n = 298)

| Factors positively associated with the birth weight |  |  |  |  |  |
| --- | --- | --- | --- | --- | --- |
| Model | Unstandardized Coefficients |  | Sig | 95.0% Confidence Interval for B |  |
|  | B | Std. Error |  | Lower | Upper |
| GAD (week) | 140.00 | 19.27 | p<0.001 | 102.07 | 177.94 |
| NANCV (unit) | 69.03 | 24.59 | .005 | 20.62 | 117.43 |
| IDDS (unit) | 97.51 | 16.97 | p<0.001 | 64.10 | 130.92 |
| Household food security status_(Food secure) | 233.20 | 54.05 | p<0.001 | 126.82 | 339.58 |
|  | Food insecure (reference category) |  |  |  |  |
| Factors negatively associated with the birth weight |  |  |  |  |  |
| Number of fetuses_(Twin) | -937.52 | 223.74 | p<0.001 | -1377.91 | -497.14 |
|  | Single (reference category) |  |  |  |  |
| Smoking habit during pregnancy_(Smoker) | -237.59 | 84.48 | .005 | -403.86 | -71.31 |
|  | Nonsmoker (reference category) |  |  |  |  |
| Pregnancy induced hypertension (Yes) | -161.25 | 68.66 | .020 | -296.39 | -26.11 |
|  | No (reference category) |  |  |  |  |

**Table 8:** Factors associated with birth weight at both host community and refugee health facilities in Gambella Region, Ethiopia, July, 2020 (n = 596)

| Factors positively associated with the birth weight |  |  |  |  |  |
| --- | --- | --- | --- | --- | --- |
| Model | Unstandardized Coefficients |  | Sig | 95.0% Confidence Interval for B |  |
|  | B | Std. Error |  | Lower | Upper |
| Household monthly income (ETB) | .01 | .00 | .010 | .00 | .02 |
| GAD (week) | 90.83 | 12.27 | p<0.001 | 66.72 | 114.94 |
| NANCV (unit) | 64.91 | 14.84 | p<0.001 | 35.77 | 94.05 |
| IDDS (unit) | 88.75 | 10.21 | p<0.001 | 68.69 | 108.81 |
| Partner educational status_(Educated) | 166.60 | 62.36 | .008 | 44.12 | 289.07 |
|  | Uneducated (reference category) |  |  |  |  |
| Maternal occupational status_(Employed) | 126.97 | 39.52 | .001 | 49.35 | 204.59 |
|  | Unemployed (reference category) |  |  |  |  |
| Household food security status_(Food secure) | 166.33 | 33.42 | p<0.001 | 100.68 | 231.97 |
|  | Food insecure (reference category) |  |  |  |  |

| Factors negatively associated with the birth weight |  |  |  |  |  |
| --- | --- | --- | --- | --- | --- |
| Number of fetuses_(Twin) | -1028.74 | 143.55 | p<0.001 | -1310.67 | -746.80 |
|  | Single (reference category) |  |  |  |  |
| Alcohol consumption during pregnancy_(Yes) | -212.58 | 49.57 | p<0.001 | -309.94 | -115.22 |
|  | No (reference category) |  |  |  |  |
| Pregnancy induced hypertension_(Yes) | -148.35 | 44.05 | .001 | -234.86 | -61.83 |
|  | No (reference category) |  |  |  |  |
| Malaria status during pregnancy_(Yes) | -81.70 | 28.01 | .004 | -136.71 | -26.68 |
|  | No (reference category) |  |  |  |  |

GAD = gestational age at delivery, NITDP = number of iron tablet during pregnancy, IDDS = individual dietary diversity score, NANCV = number of antenatal care visits

## Discussion

Birth weight is a major indicator that determines child’s probability to survive as well as susceptibility to various risks of childhood morbidities later in life. In this study, the mean birth weight of neonates in host community was found to be 3282.55±415.97 grams. This finding is consistent with the study done in Jimma (3245.3±51.8 grams) (15). This might be due to similarity in the background of the study participants. However, this result is slightly difference with the study done in Bahir Dar (2707.7±518.4 grams) (19) and South Africa (2675.48±616.16 grams) (20). This difference might be due to design, time and area difference with the current study. On the other hand the study found the neonatal birth weight in the refugee to be 3109.40±635.10 grams which is also consistent with the study done in South Sudan (3100±600 grams) (17). This consistence might also be due to similarity in the background of the study participants. Moreover, the current study found statistical significant difference between the mean birth weights of host and refugee neonates. This finding is consistent with the study done in Portugal with mean birth weights of 3303±424 and 3297±441 grams among host and refugee neonates respectively (p<0.05) (21). This consistence might be due to similarity in the designs, inclusion of sufficient number of respondents and common backgrounds of the study participants in both studies. Moreover, the larger mean birth weight among host neonates might generally reflect the fact that host people meet more basic and necessary needs for the health life as they are staying in their own residences compared to the refugee counterpart.

The current study found that number of fetuses is statistically significantly associated with the birth weight. This finding is consistent with the study done in Ethiopia using EDHS data (22). This consistence might be due to the fact that the fetuses share the same uterus during their development. Gestational age was found to be associated with the birth weight and that when gestational age increases, the birth weight goes on increasing. This finding is consistent with the study done in Bahir Dar (19), Hawassa (23), Gonder (24) and Kenya (25). This consistence might be due to the fact that increases in gestational age leads the fetus to fully develop which in turn results in increment of the neonatal birth weight. Household food security status was significantly associated with the birth weight in the current study that is mothers in food secure household have less chance of delivering low birth weight. This is supported by the finding of the case control study done in Addis Ababa (26). This might be due to the fact that mothers in household experiencing food security may increase food intake, consume a good quality food and always conserve their daily meal patterns. This study showed that individual dietary diversity score is associated with the birth weight and that when the individual dietary diversity score increases, the birth weight also increases. This is consistent with the study done in Dessie town (27). This consistence might be due to improvement of the nutritional status of not just the mother but her neonate as well which results in an increase of the birth weight as the dietary diversity of fetus depends on that of the mother. Hence, the more the diversify the maternal diets are the more the dietary diversity of the fetus is likely to be so that birth weight will become higher as a result of dietary improvement.

Pregnancy induced hypertension was another factor significantly associated with the birth weight in the current finding and that mother who had pregnancy induced hypertension are more likely to deliver low birth weight baby compared to those who had not. This finding is consistent with the study done in Gonder (24) and Addis Ababa (26). This might be due to the reduction in the delivery of nitrogen and oxygen to the placenta as a result of the decrease of the blood flow through spiral artery so the birth weight of the neonates will become lower as a result.

### Strength of the study

This study is the first to be conducted in the area. Data were gathered within 24 hours after birth which possibly may minimize the recall bias. Being a comparative study in nature is one of the added strengths of the study as it provide evident in both the areas at the same time. Unlike many, the sample size included for this study is large and that it has 100% of response rate which may maximize the precision of the study.

### Limitation of the study

Despite its strengths, the study was conducted in the health facility alone in its scope excluding the home delivered mothers and a cross-sectional in nature, so generalization to all mothers in the area might be in doubt and might not capture the seasonal variations. A certain level of recall bias was expected on some variables specially, on menstrual, food security and dietary related habits. Some important factors such as BMI, weight gained during pregnancy, MUAC as well as household, environmental and cultural related factors were not included in addition to gathering the birth weight data directly from the health institutional records which might lack appropriate control measure.

### Conclusions

The mean birth weights in the two settings were found to be significantly different. Host community neonates have larger mean birth weight compared to that of refugee neonates at health facilities in Gambella Region.

Many factors were identified to be significantly associated with the birth weight. Among those significant factors, five of them were identified as the most important factors significantly associated with the birth because of their influent on the outcome variable.

The five most important factors for the birth weight in this study are, number of fetuses, gestational age at delivery, house hold food security, individual dietary diversity score and pregnancy induced hypertension. These factors play crucial role in the region as they are significantly associated with the birth weight not just in host or refugee alone but in each setting and on the combined analysis (region) of both host and refugee as well.

## Data Availability

The datasets used and/or analysed during the current study are available from the corresponding author on reasonable request

NA

## Declaration

### Ethics approval and consent to participate

This study received ethics approval from Ethical Review Committee of College of Medicine and Health Sciences of Wollo University (ID: CMHS1198/13/12)

### Consent for publication

All health facilities were informed about the purpose of the study in the local language, had the opportunity to ask questions and then consented and in addition verbal consent was received from each study participant.

### Authors’ contributions

**BCN** developed the proposal, designed the study, monitored the data collection, performed the statistical analysis and drafted the final paper. **SW and SET** critically reviewed the paper, gave the guidance, gave advices and approved the proposal and the final paper. All authors read and approved the final manuscript.

## Acknowledgement

We are so much grateful to Wollo University for the approval of the ethical clearance and to Gambella University for the financial support for this study. Furthermore, we would like to thanks all data collectors and supervisors for their extraordinary contribution on this study. We would also like to thanks all study participants for taking their time participating in this research.

